# Prime hAd5 Spike + Nucleocapsid Vaccination Induces Ten-Fold Increases in Mean T-Cell Responses in Phase 1 Subjects that are Sustained Against Spike Variants

**DOI:** 10.1101/2021.04.05.21254940

**Authors:** Peter Sieling, Thomas King, Raymond Wong, Andy Nguyen, Kamil Wnuk, Elizabeth Gabitzsch, Adrian Rice, Helty Adisetiyo, Melanie Hermreck, Mohit Verma, Lise Zakin, Annie Shin, Brett Morimoto, Wendy Higashide, Kyle Dinkins, Joseph Balint, Victor Peykov, Justin Taft, Roosheel Patel, Sofija Buta, Marta Martin-Fernandez, Dusan Bogunovic, Patricia Spilman, Lennie Sender, Sandeep Reddy, Philip Robinson, Shahrooz Rabizadeh, Kayvan Niazi, Patrick Soon-Shiong

## Abstract

In response to the need for a safe, efficacious vaccine that elicits vigorous T cell as well as humoral protection against SARS-CoV-2 infection, we have developed a dual-antigen COVID-19 vaccine comprising both the viral spike (S) protein modified to increase cell-surface expression (S-Fusion) and nucleocapsid (N) protein with an Enhanced T-cell Stimulation Domain (N-ETSD) to enhance MHC class I and II presentation and T-cell responses. The antigens are delivered using a human adenovirus serotype 5 (hAd5) platform with E1, E2b, and E3 regions deleted that has been shown previously in cancer vaccine studies to be safe and effective in the presence of pre-existing hAd5 immunity. The findings reported here are focused on human T-cell responses due to the likelihood that such responses will sustain efficacy against emerging variants, a hypothesis supported by our *in silico* prediction of T-cell epitope HLA binding for both the first-wave SARS-CoV-2 ‘A’ strain and the B.1.351 strain K417N, E484K, and N501Y spike and T201I N variants. We demonstrate the hAd5 S-Fusion + N-ETSD vaccine antigens expressed by previously SARS-CoV-2-infected patient dendritic cells elicit Th1 dominant activation of autologous patient T cells, indicating the vaccine antigens have the potential to elicit immune responses in previously infected patients. For participants in our open-label Phase 1b study of the vaccine (NCT04591717; https://clinicaltrials.gov/ct2/show/NCT04591717), the magnitude of Th-1 dominant S- and N-specific T-cell responses after a single prime subcutaneous injection were comparable to T-cell responses from previously infected patients. Furthermore, vaccinated participant T-cell responses to S were similar for A strain S and a series of spike variant peptides, including S variants in the B.1.1.7 and B.1.351 strains. The findings that this dual-antigen vaccine elicits SARS-CoV-2-relevant T-cell responses and that such cell-mediated protection is likely to be sustained against emerging variants supports the testing of this vaccine as a universal booster that would enhance and broaden existing immune protection conferred by currently approved S-based vaccines.

## INTRODUCTION

To address the need for an efficacious COVID-19 vaccine that elicits broad immunity against SARS-CoV-2 and has a high likelihood of maintaining efficacy against emerging SARS-CoV-2 variants, we have developed a dual-antigen COVID-19 ‘T-cell’ vaccine. This dual-antigen vaccine expresses both the virus spike (S) and nucleocapsid (N) proteins using a next-generation human adenovirus serotype 5 (Ad5) platform. The S antigen is full-length S including SD1 receptor binding domain, S1 and S2 domains modified to enhance surface expression (S-Fusion); ^1^ and full-length N protein modified with an Enhanced T-cell Stimulation Domain (ETSD) to direct N to the endo/lysosomal compartment for increased MHC class I and II expression. The hAd5 platform has deletions in the E1, E2b, and E3 gene regions, thereby minimizing host anti-vector immune responses ^2,3^ and enabling efficient antigen cargo expression and cognate T-cell activation even in the presence of existing anti-adenovirus immunity, as demonstrated previously in clinical studies targeting tumor-associated antigens in cancer patients. ^4,5^

The emergence of SARS-CoV-2 variants that have the potential to evade immune responses generated in response to currently available vaccines has spurred renewed interest in the potential role of T cells in conferring long-term protection against COVID-19. ^6-9^ SARS-CoV-2 vaccine development has focused largely on eliciting neutralizing antibodies against the S protein to inhibit infection and reduce disease severity, ^10-14^ however, S-specific T-cell responses have been reported for two available vaccines, mRNA1273 and BNT162b1. ^15,16^

SARS-CoV-2 strain analysis in participants in Phase 3 studies in South Africa has revealed that some vaccines show diminished protection against the B.1.351 variant strain, ^17^ including ChAdOx, ^18^ NVX-CoV2373, ^19,20^ and Ad26.COV2-S. ^21^ In addition, *in vitro* studies show that wild type S-specific antibodies elicited by mRNA-1273 and BNT162b1 show reduced binding to the B.1.351 variant S protein. ^22-25^ Taken together, these findings raise the concern that monovalent vaccines targeting only the S protein may not be an optimal strategy for conferring protection against continually emerging variants.

SARS-CoV-2 expresses a number of other immunogenic proteins that may induce protective antibody and/or T-cell responses. Among these, the N protein is the most abundantly expressed, is highly conserved among coronaviruses, and has been studied previously as an antigen for therapeutics and vaccines for SARS-CoV.^26-30^ Inclusion of N-derived antigens into vaccine designs may be a rational strategy to elicit broader protective immunity against SARS-CoV-2 with a high probability of being sustained against variants. The N protein associates with viral RNA within the SARS-CoV-2 virion and is necessary for viral RNA replication, virion assembly, and release from host cells.^29,30^ Virtually all SARS-CoV-2 convalescents develop N-specific antibody and CD8^+^ and/or CD4^+^ T-cell responses.^31^ SARS-CoV convalescents from 2003 maintain robust cognate N-specific CD8^+^ and/or CD4^+^ T-cell responses that were detectable up to 17 years later.^32^ These long-lived memory T cells also cross-react with N and other proteins of SARS-CoV-2.^32^ To a lesser extent, individuals with no history of exposure to SARS-CoV, SARS-CoV-2, or Middle Eastern Respiratory Syndrome (MERS) also show cross-reactive SARS-CoV-2 N-specific CD8^+^ and/or CD4^+^ T-cell responses, likely due to prior exposure to related endemic coronaviruses including OC43, HKU1, 229E, and NL63.^31^ These data suggest that induction of N-specific memory T-cell responses via vaccination may at least partly recapitulate the natural course of SARS-CoV-2 infection and recovery from illness, and could be beneficial for achieving more optimal disease-limiting immunity against the virus.

Recent characterization of COVID-19 patient immune responses to SARS-CoV-2 indicates T-cell activation is critical for clearance of infection and production of long-term immunity to coronavirus infections.^32-35^ Both CD4+ and CD8+ T cells underpin durable immune responses because CD4+ T cells, while not effector cells like CD8+ T cells, are critical to the generation of robust and long-lasting immunity afforded by antibody-secreting plasma cells and the elimination of infected cells by memory cytotoxic CD8+ T cells.^36,37^

Here, to support our hypothesis that a vaccine that generates vigorous T-cell responses has a reasonable likelihood of sustaining efficacy against emerging variants, we first describe our *in silico* identification of 9-mer T-cell epitopes and prediction of HLA binding for both first-wave ‘A’ strain SARS-CoV-2 and the variants that are found in the B.1.351 strain discovered in South Africa – spike K417N, E484K, and N501Y and nucleocapsid T205I.

We then demonstrate that the hAd5 S-Fusion + N-ETSD (hAd5 S + N) COVID-19 vaccine antigens, when expressed by monocyte-derived dendritic cells (MoDCs) from individuals previously infected with SARS-CoV-2, can activate and induce recall of S- and N-specific autologous CD8^+^ and CD4^+^ T cells. Overall, N-ETSD was found to elicit greater T-cell responses than N without the ETSD. This study demonstrates the vaccine-expressed antigens are ‘recalled’ by T cells and thus suggests they have the potential to induce relevant, protective immune responses in previously SARS-CoV-2 infected or vaccinated individuals who receive the hAd5 S + N vaccine.

We further report that T cells isolated 14 and 23 days after prime vaccination alone from participants in our ongoing Phase 1b clinical trials of the hAd5 S + N vaccine are activated by both S and N peptides pools and that these T-cell responses are not only comparable in magnitude to those of T cells from previously SARS-CoV-2 infected patients, activation by S peptides is sustained for variant S peptides, including those found in the B.1.1.7, B.1.429, P.1 and B.1.351 strains.

## MATERIALS and METHODS

### *In silico* T-Cell Epitope HLA Binding Prediction

#### SARS-CoV-2 reference sequences

The reference strain and COVID19 variants used for epitope analysis was assembled from 75,000 individual sequences deposited in the GISAID database. Binding analysis was run on the consensus sequence generated using our in-house binding algorithm.^38^ Briefly, neural network classifiers that process full HLA protein and query peptide amino acid sequences were trained on experimental binding data deposited in the IEDB database.^39^ A classifier ensemble was leveraged to abstain from making predictions when output uncertainty overlapped the decision threshold, thus boosting robustness and precision of binding prediction results.

The variants assessed included (name/mutation(s)): B.1.1.7/A222V mutation; B.1.1.7 /N501Y mutation; B.1.351 and B.1.1.28/K417N (or K417T for B.1.1.28), E484K, and the N501Y mutations; and P.1/E484K. Binding efficiencies as well as HLA-epitope pairs were compared between reference sequences and mutations to determine potential immune evasion due to the reported variants.

### Production of hAd5 S + N and related SARS-CoV-2 antigen expressing vectors

Production of all hAd5 [E1-, E2b-, E3-] (Fig. 1A) constructs and virus particles were carried out as previously described.^40^ In brief, high titer adenoviral stocks were generated by serial propagation in the E1- and E2b-expressing E.C7 packaging cell line, followed by CsCl2 purification, and dialysis into storage buffer (2.5% glycerol, 20 mM Tris pH 8, 25 mM NaCl) by ViraQuest Inc. (North Liberty, IA). Viral particle counts were determined by sodium dodecyl sulfate disruption and spectrophotometry at 260 and 280 nm. Viral titers were determined using the Adeno-X™ Rapid Titer Kit (Takara Bio).

**Fig. 1.**
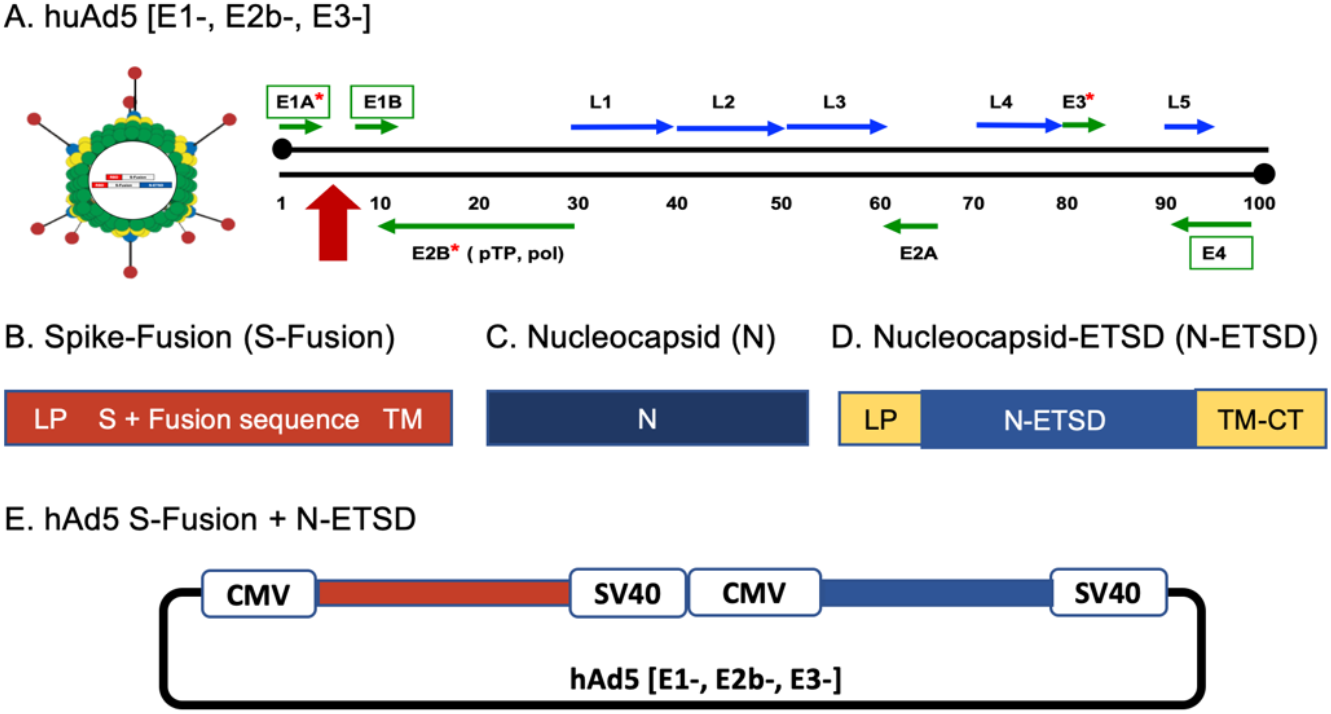
Human adenovirus serotype 5 (hAd5) platform and constructs. (A) hAd5 with E1, E2b and E3 regions deleted (hAd5[E1-, E2b-, E3-]). The site of antigen sequence insertion is shown with a red arrow. Constructs tested included (B) spike (S)-Fusion, (C) nucleocapsid (N) without the Enhanced T-cell Stimulation Domain (ETSD), (D) N-ETSD, and (E) the hAd5 S-Fusion + N-ETSD vaccine.

The constructs created included:

Fig. 1B. S-Fusion: S optimized to enhance surface expression and display of RBD.

Fig. 1C. N (N without ETSD): Nucleocapsid (wild type) sequences with tags for immune detection, but without ETSD modification, and predominantly cytoplasmic localization.

Fig. 1D. N with the Enhanced T-cell Stimulation Domain (N-ETSD): Nucleocapsid (wild type) with ETSD to direct endo/lysosomal localization and tags for immune detection.

Fig. 1E. The dual-antigen hAd5 S-Fusion + N-ETSD vaccine.

### The T-cell recall studies of unexposed and previously SARS-CoV-2 infected individuals

#### Collection of plasma and PBMCs for differentiation of MoDCs from patients with confirmed previous SARS-CoV-2 infection and from virus-naïve volunteers

Blood was collected with informed consent via venipuncture from volunteer patients (Pt) who had either not been exposed (UNEX) to SARS-CoV-2 as confirmed by ELISA and multiple negative SARS-CoV-2 tests or who had recovered from COVID-19 as indicated by recent medical history and a positive SARS-CoV-2 antibody test. The presence of anti-S IgG and of neutralizing antibodies in plasma from previously SARS-CoV-2 infected patients was confirmed by ELISA, a surrogate neutralization assay,^41^ and a live virus assay as shown in Supplementary Fig. S1. A third source of whole blood was apheresis of healthy subjects from a commercial source (HemaCare).

Peripheral blood mononuclear cells (PBMCs) were isolated from whole blood by density gradient centrifugation and plasma was collected after density gradient centrifugation.

Monocyte-derived dendritic cells (MoDC) were differentiated from PBMC using GM-CSF (200U/ml) and IL-4 (100U/ml) as previously described ^42^. Briefly, monocytes were enriched by adherence on plastic, while the non-adherent cells were saved and frozen as a source of lymphocytes, specifically T cells. Adherent cells were differentiated into dendritic cells (3-5 d in RPMI containing 10% FBS), then frozen in liquid nitrogen for later use. T cells were enriched from the non-adherent fraction of PBMC using MojoSort (BioLegend CD3 enrichment). CD4+ and CD8+ T cells were enriched using analogous kits from the same manufacturer. Efficiency of the cell separations was evaluated by flow cytometry.

#### Transduction of MoDCs with hAd5 N-WT or N-ETSD and labeling with anti-N, anti-CD71, anti-LAMP-1, and Anti-LC3a/b antibodies

Freshly thawed MoDCs were plated on 4-well Lab-Tek II CC2 Chamber Slides, using 3 × 10^4^ cells per well and transduction performed at MOI 5000 one hour after plating using hAd5 N-ETSD or hAd5 N. Slides were incubated o/n at 37°C, fixed in 4% paraformaldehyde for 15 minutes, then permeabilized with 1% Triton X100, in PBS) for 15 min. at room temperature. To label N, cells were then incubated with an anti-flag monoclonal (Anti-Flag M2 produced in mouse) antibody at 1:1000 in phosphate buffered saline (PBS) with 3% BSA, 0.5% Triton X100 and 0.01% saponin overnight at 4°C, followed by three washes in PBS and a 1 hour incubation with a goat anti-Mouse IgG (H+L) Highly Cross-Adsorbed Secondary Antibody, Alexa Fluor Plus 555 (Life Technologies) at 1:500. For co-localization studies, cells were also incubated overnight at 4°C with a rabbit anti-CD71 (transferrin receptor, recycling/sorting endosomal marker) antibody (ThermoFisher) at 1:200; sheep anti-Lamp1 Alexa Fluor 488-conjugated (lysosomal marker) antibody (R&D systems) at 1:10; or a rabbit monoclonal anti human LC3a/b (Light Chain 3, autophagy marker) antibody (Cell Signaling Tech #12741S) used at 1:100. After removal of the primary antibody, two washes in PBS and three washes in PBS with 3% BSA, cells were incubated with fluor-conjugated secondary antibodies when applicable at 1:500 (Goat anti-Rabbit IgG (H+L) secondary antibody, Alexa Fluor 488; 1:500 dilution) for 1 hour at room temperature. After brief washing, cells were mounted with Vectashield Antifade mounting medium with DAPI (Fisher Scientific) and immediately imaged using a Keyence all-in-one Fluorescence microscope camera and Keyence software.

#### CD3+ T-cell and selected CD4+ and CD8+ T-cell secretion of IFN-γ in response to MoDCs pulsed with SARS-CoV-2 peptide antigen pools

The ability of T cells from previously infected patients used in these studies to recognize SARS-CoV-2 antigens *in vitro* was validated and then similar analyses were performed for selected CD4+ and CD8+ T cells. Briefly, MoDC (2 × 10^4^) were pulsed with SARS-CoV-2 peptide antigens (1μg/ml, PepMix S comprising the S1 and S2 pools PM-WCPV-S-1; and N PM-WCPV-NCAP-1, both JPT Peptide Technologies) then autologous T cells (1 × 10^5^), enriched from the non-adherent fraction of PBMC using MojoSort (BioLegend CD3 enrichment) were added in enriched RPMI (10% human AB serum). Cells were cultured in a microtiter plate (Millipore) containing an immobilized primary antibody to target IFN-γ, overnight (37°C), then IFN-γ spot forming cells enumerated by ELISpot. For ELISpot detection, after aspiration and washing to remove cells and media, IFN-γ was detected by a secondary antibody to cytokine conjugated to biotin. A streptavidin/horseradish peroxidase conjugate was used detect the biotin-conjugated secondary antibody. The number of spots per well (1 × 10^5^ cells), was counted using an ELISpot plate reader. IL-4 was measured by ELISpot using a kit (MabTech) with wells precoated with anti-IL-4 antibody and following the manufacturer’s instructions. Remaining steps for IL-4 detection were identical to those for IFN-γ, but with alkaline phosphatase detection rather than peroxidase.

#### Determination of previously SARS-CoV-2 infected patient-derived T-cell reactivity in response to autologous hAd5 vaccine-transduced MoDCs

MoDCs were transduced with hAd5 S-Fusion, S-Fusion + N-ETSD, N-ETSD, N or GFP/Null constructs and incubated overnight at 37°C. The transduced MoDCs were cultured with CD3+, CD4+, or CD8+ T cells from the same individuals overnight. Antigen specific T-cell responses were enumerated using ELISpot as described above.

### Phase 1b Clinical Trial and Analyses

#### Vaccination of healthy adult subjects with hAd5 S + N

The current Good Manufacturing Practice (cGMP) vaccine used in the clinical trial was prepared using the generated high titer adenoviral stocks described above in *Production of hAd5 S + N and related SARS-CoV-2 antigen expressing vectors*.

In a phase 1b, open-label study (QUILT 4.001 in adults volunteers conducted at Hoag Hospital in Orange County, California, USA; the hAd5 S + N vaccine was administered subcutaneously (SC) at a dosage of 1×10^11^ VP to cohort 2 participants.

Study participants comprised healthy adult volunteers between the ages of 18 and 55. Subjects were assigned to cohorts. Data from cohort 2 (high dose) is presented here. All the participants provided informed, written consent before enrollment.

#### Trial Oversight

The trial protocol was approved by the Western IRB for the Chan Soon-Shiong Institute for Medicine (CSSIFM) and the Hoag Hospital Providence IRB at Hoag Hospital Newport Beach. ImmunityBio, Inc. was the regulatory sponsor of the trial and holder of the Investigational New Drug (IND) application. The trial was funded by ImmunityBio, Inc. The vaccine was designed and manufactured by ImmunityBio. *The studies presented here are experimental and all assays and analyses were performed in-house at ImmunityBio, Inc. Formal analyses of the primary and secondary end points are not reported in this manuscript*.

#### T-cell analyses

T-cell responses in PBMCs were measured at baseline (Day 1) and after prime vaccination alone on Day 14-16 and 21-23 by ELISpot for secretion of interferon γ (IFNγ) and interleukin 4 (IFN-4). T helper cell type 1 dominance was determined based on the IFNγ/IL-4 ratio.

#### IFN-γ ELISpot for enumeration of S- and N-specific T cells in PBMC

ELISpot assays were used to enumerate S- and N-specific IFN-γ-secreting T cells in fresh PBMCs isolated from the blood of subjects vaccinated with hAd5 S + N. Whole blood was collected by venipuncture pre- and post-vaccination. PBMCs were isolated from whole blood by standard density gradient centrifugation and frozen in liquid nitrogen until use. On the day of assay, PBMCs were thawed and re-suspended in RPMI 10% human AB serum, then incubated with 2.5 μg/ml of SARS-CoV-2 S or N peptide pools (JPT Peptide Technologies catalogue # PM-WCPV-S and PM-WCPV-NCAP-1, respectively). Internal positive controls were incubated with 2.5 μg/ml of CEFT peptide pool consisting of epitopes derived from human Cytomegalovirus, Epstein-Barr virus, Influenza A, and Clostridium tetani (JPT Peptide Technologies catalogue # PM-CEFT-4). Positive assay controls were incubated with a CD3/CD28/CD2 agonist (StemCell Technologies catalogue # 10970). Negative controls were incubated with media alone or 2 μg/ml of SARS-CoV-2 membrane peptide pool (JPT Peptide Technologies catalogue # PM-WCPV-VME). The number of spot-forming cells (SFCs) per well (2.5-4 × 10^5^ PBMCs) were enumerated using an ELISpot plate reader (Cellular Technologies Limited model # S6UNIV-01-7115). All IFN-γ ELISpot reagents were obtained from a commercial kit and used according to manufacturer protocol (Cellular Technologies Limited catalogue # Hu IFN-g/IL-4).

#### Variant S peptides

For the assessment of PBMC responses to variant S peptides, ELISpot assays were performed as described above using peptide mixes from JPT. Each mix comprised 315 peptides delivered in 2 subpools of 158 and 157 peptides derived from a peptide scan through the entire spike glycoprotein of SARS-CoV-2. The mixes included: PepMix™ SARS-CoV-2 lineage B.1.1.7 (United Kingdom; PM-SARS2-SMUT01-1) covering the following mutations: H0069-, V0070-, Y0144-, N0501Y, A0570D, D0614G, P0681H, T0716I, S0982A, D1118H; PepMix™ SARS-CoV-2 lineage B.1.351 (South Africa; PM-SARS2-SMUT02-1) covering the following mutations: D0080A, D0215G, L0242-, A0243-, L0244-, K0417N, E0484K, N0501Y, D614G, A0701V; PepMix™ SARS-CoV-2 lineage P.1 (Brazil; PM-SARS2-SMUT03-1) covering the following mutations: L0018F, T0020N, P0026S, D0138Y, R0190S, K0417T, E0484K, N0501Y, D0614G, H0655Y, T1027I, V1176F; and PepMix™ SARS-CoV-2 lineage B.1.429 (Los Angeles; PM-SARS2-SMUT04-1) covering the following mutations: S0013I, W0152C, L0452R, D0614G.

## RESULTS

### T-cell epitope HLA binding prediction

#### In silico identification of 9-mer T-cell epitopes and HLA binding prediction suggests such binding is not significantly affected by B.1.351 strain variants

To understand and predict the possible effects emerging SARS-CoV-2 variants may have on vaccine-elicited T-cell protection, we utilized our in-house T-cell epitope identification and binding analysis pipeline to compare the first wave ‘A’ strain epitopes to the B.1.351 variant strain.

*In silico* epitope analysis of the reference strain assembled from 75,000 individual sequences deposited in the GISAID database ^43,52^ revealed 2479 MHC class I and 20963 MHC class II potential binders. Altogether, the SARS-CoV-2 variants (K417N, E484K and N501Y mutations) of the B.1.351 strain generate 10970 MHC class I and 18017 MHC class II unique epitopes.

We compared these unique epitopes to the reference strain and counted the different epitopes that are bound by HLAs, finding 1659 unique MHC class I and 2887 unique MHC class II epitopes that only exist in the variants and not the original strain. These epitopes are from the S, envelope (E), N and membrane (M) proteins, but the most unique epitopes were found in variant S due to the higher numbers of coding mutations.

For HLA binding prediction, sequences and predicted binding were compared and aligned, and we found that, for example, as compared to the ‘A’ strain, a B.1.351 sequence has a 3 amino acid deletion that results in a unique sequence RFQTLHRSYL predicted to bind HLA-C*14:02. By similar determination of the epitopes and HLAs from the reference strain and the B.1.351 variants, differences were identified in binding affinity or HLAs. Similarly at the N501Y site, the reference strain generates an epitope (TNGVGYQPY) predicted to bind HLA-A*30:02, while N501Y would generate a different yet similar epitope (TYGVGYQPY) also predicted to bind HLA-A*30:02.

The key questions asked here were: how does mutation change HLA availability and which mutations favor immune evasion? Not all mutations lead to evasion; while a mutation may lead to an amino acid substitution that has potential functional consequences, that same mutation may still generate an epitope that retains HLA binding affinity and does not contribute to escape. The unique binding HLAs for each B.1.351 S variant when compared to the A strain are shown in Figure 2. For class I, for K417N there are only 2 unique HLAs and for the A strain there are 6 (Fig. 2A); for E484K there are 8 unique HLAs and for the A strain 7 (Fig. 2B); and for N501Y there are 8 unique HLAs and for the A strain only 1 (Fig. 2C). There are no unique class II HLAs for these same comparisons (Fig. 2D-F).

**Fig. 2.**
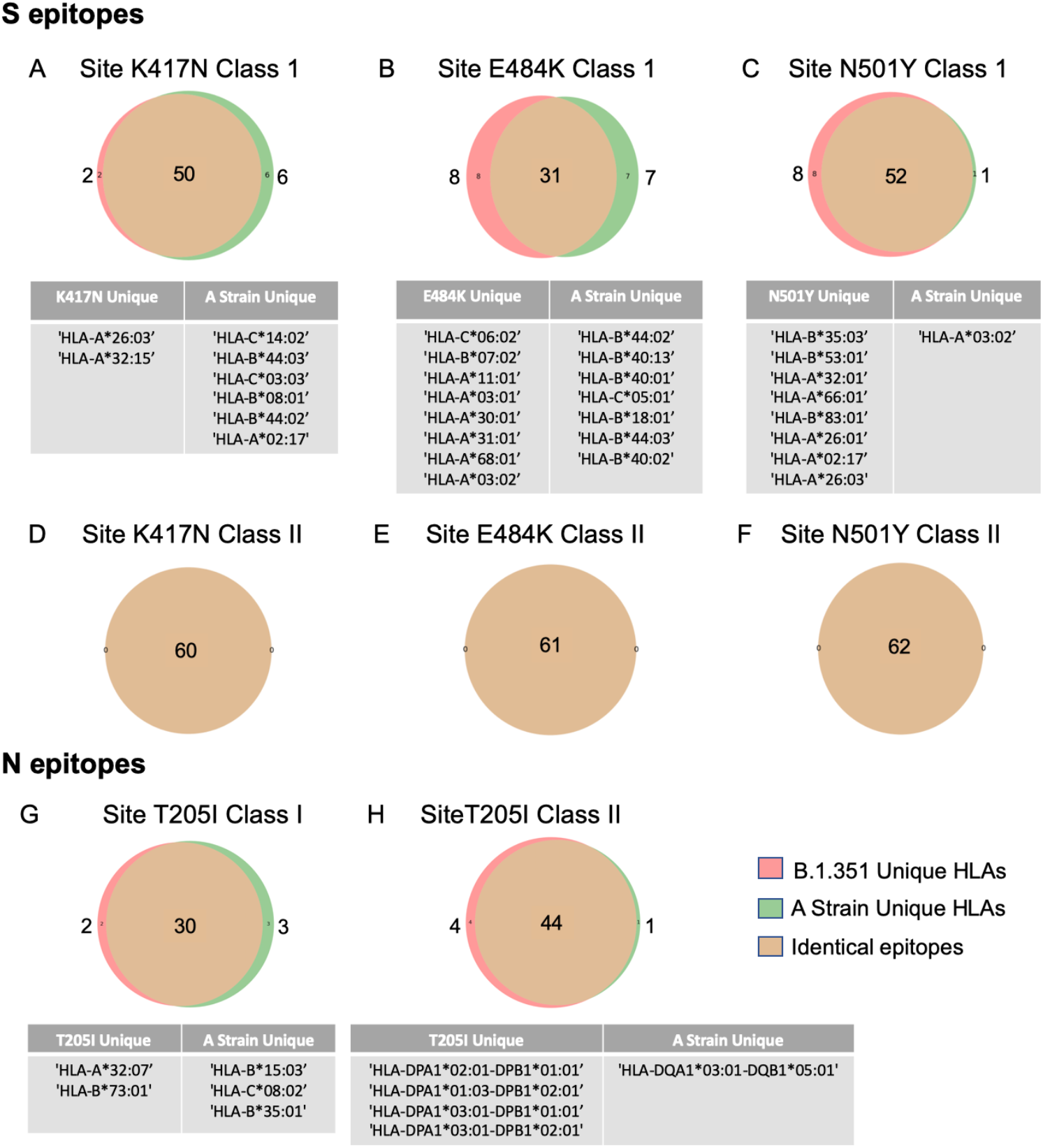
A high degree of overlap is seen for HLA binding of predicted T-cell epitopes from variants and the first-wave A strain SARS-CoV-2. Comparison of ‘A’ and B.1.351 strain predicted class I T-cell epitopes reveals significant overlap for the spike (A) K417N (50 of 58 epitopes), (B) E484K (31 of 46 epitopes), and (C) N501Y (52 of 61 epitopes) variants. (D-F) Complete overlap is seen for class II epitopes for those variants, respectively. (G) For T205I class I, 30 of 35 epitopes overlap and (H) for class II, 44 of 49. Unique HLA class I binding is shown in the tables.

In the B.1.351 strain, there is only one N mutation – T205I. Similarly to the S variants, there are few unique binding HLAs. For class I, the T201I mutation generates 2 and the A strain 3 unique binding HLAs (Fig. 2G). For class II, the T205I generates 4 and the A strain just 1 unique binding HLA(s) (Fig. 2H).

### N-ETSD is directed to endo/lysosomal and autophagosomal compartments

The hAd5 dual antigen vaccine construct includes sequences designed to target N to MHC class I and II antigen loading compartments (endo/lysosomes) to enhance T-cell responses. Wildtype S is a surface-expressed protein, therefore the addition of a targeting sequence is not necessary. To confirm N-ETSD is directed to endo/lysosomes, we evaluated the localization of N-ETSD in human monocyte-derived dendritic cells (MoDCs) compared to its untargeted cytoplasmic counterpart (N). MoDCs from healthy subjects were infected with hAd5 N-ETSD or hAd5 N and localization was determined by immunocytochemistry. N-ETSD showed localization to discrete vesicles, some coincident with CD71, a marker of recycling endosomes (Fig. 3A-C), and LAMP-1, a marker for late endosomes/lysosomes (Fig. 3G-I), whereas untargeted N was expressed diffusely and uniformly throughout the cytoplasm (Fig. 3D-F; J-L).

**Fig. 3.**
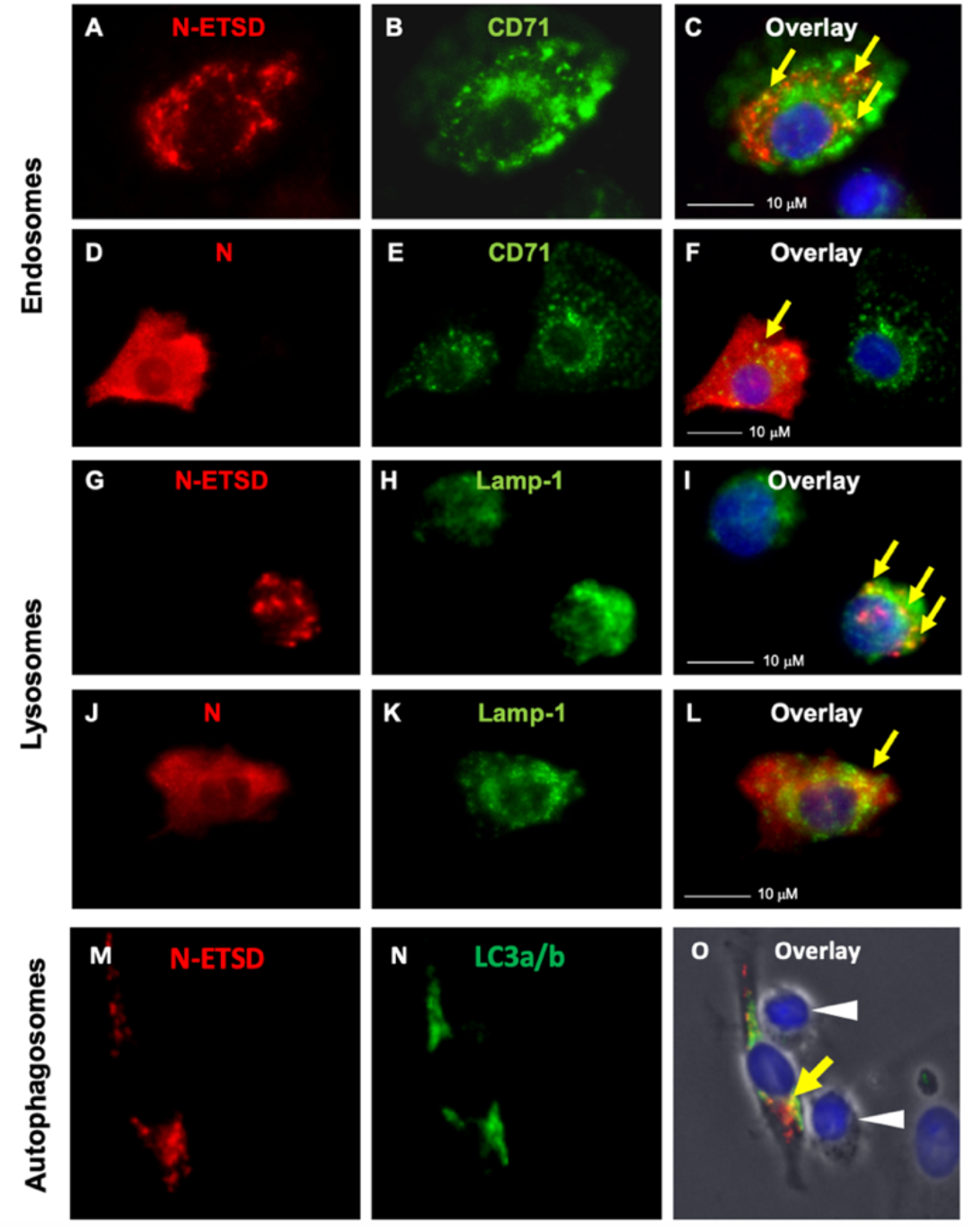
N-ETSD localizes to endosomes, lysosomes, and autophagosomes. MoDCs were infected with Ad5 N-ETSD or N without ETSD and were co-labeled with anti-flag (N, N-ETSD here have a flag tag) and anti-CD71 (endosomal marker), anti-Lamp 1 (lysosomal marker), or anti-LC3a/b antibodies. (A) N-ETSD, (B) CD71, and (C) overlay. (D) N, (E) CD71, and (F) overlay. (G) N-ETSD, (H) Lamp-1, and (I) overlay. (J) N, (K) Lamp-1, and (L) overlay. (M) N-ETSD, (N) LC3a/b, and (O) overlay. N/N-ETSD is red, other markers green, co-localization indicated by yellow arrows, and white arrows indicate suspected undifferentiated lymphocytes.

Studies have shown that lysosomes fuse with autophagosomes to enhance peptide processing and MHC class II presentation.^44,45^ Thus we examined whether N-ETSD localized in autophagosomes in MoDCs by co-labeling with the autophagosome marker LC3a/b ^46^ to identify another potential site of localization relevant to MHC class II antigen presentation.^47^ We found that N-ETSD also displayed some co-localization with the autophagosome marker (Fig. 3M-O).

Protein processing in autophagosomes plays a key role in MHC-mediated antigen presentation in DCs,^48-50^ providing a potential mechanism of enhanced CD4+ T cells induced by N-ETSD in the vaccine construct. Evidence of this T-cell interaction with a MoDC infected with N-ETSD translocated to autophagosomes (and, it is assumed, also endosomes and lysosomes) is seen in this phase-contrast microscopy of the N-ETSD and LC3a/b co-labeled cells, which reveals the elongated DC morphology in contrast to the spherical morphology of what are posited to be undifferentiated lymphocytes.

### T-Cell memory in patients previously infected with SARS-CoV-2

#### Blood samples from previously SARS-CoV-2 infected patients show antibody and T-cell responses and enhanced binding to hAd5 S + N expressing cells

For the ‘recall’ studies described below, plasma samples were collected from four individuals convalescing from SARS-CoV-2 infection as confirmed by antibody assays and patient history. The presence of anti-Spike IgG, and neutralizing antibodies by both a surrogate SARS-CoV-2 neutralization ^41^ and live virus assays, were confirmed in all patient samples (Supplementary Fig. S1). Samples from four virus-naïve individuals were used as controls.

In additional studies to validate immune responses to SARS-CoV-2 antigens, the binding of previously SARS-CoV-2 infected patient and virus-naïve control individual plasma to human embryonic kidney (HEK) 293T cells transfected with either hAd5 S-Fusion alone or hAd5 S + N was assessed (Supplementary Fig. S2). This binding reflects the presence of antibodies in plasma that recognize antigens expressed by the hAd5 vectored vaccines. Quantification of histograms showed little or no binding of virus-naïve plasma antibodies to cells expressing constructs, with the highest previously SARS-CoV-2 infected patient plasma binding to cells expressing the dual antigen S-Fusion + N-ETSD construct (Supplementary Fig. S2R).

To confirm the T cells from previously SARS-CoV-2-infected patients were reactive to SARS-CoV-2 peptides, isolated T cells from the patients were incubated with S1 (containing the S receptor binding domain), S2 and N peptides pools. T cells from all four patients showed reactivity (Supplementary Fig. S3). Selected CD4+ and CD8+ T cells from 2 patients were activated by S1, S2, and N peptides, revealing CD8+ T cells were more reactive to N (Supplementary Fig. S3D and E).

#### Previously SARS-CoV-2 infected patient Th1 dominant CD3+ and CD4+/CD8+ memory T-cells recall N-ETSD antigens expressed by autologous hAd5-infected MoDCs

To evaluate the immune significance of endo/lysosome-localized N-ETSD versus cytoplasmic N, MoDCs were infected with hAd5 Null, N-ETSD or N constructs then incubated with autologous CD3+-, or CD4+-/CD8+-selected T cells (Fig. 4A). CD3+ T cells from previously infected SARS-CoV-2 patients showed significantly greater IFN-γ secretion in response to N-ETSD than both Null and cytoplasmic N in the two patients (Pt 3 and Pt 1) where N-ETSD and N were compared (Fig. 4F and J). Interleukin-4 (IL-4) secretion by CD3+ T cells was low for all patients (Fig. 4C, G, and K); high IFN-γ and low IL-4 responses show Th1 dominance for both N-ETSD and N. Both CD4+ and CD8+ selected T-cell populations showed significantly greater IFN-γ responses to N-ETSD than Null (Fig. 4D, E H, I, L and M) and in two of three patients, CD4+ and CD8+ T cells showed greater recognition of N-ETSD compared to N.

**Fig. 4.**
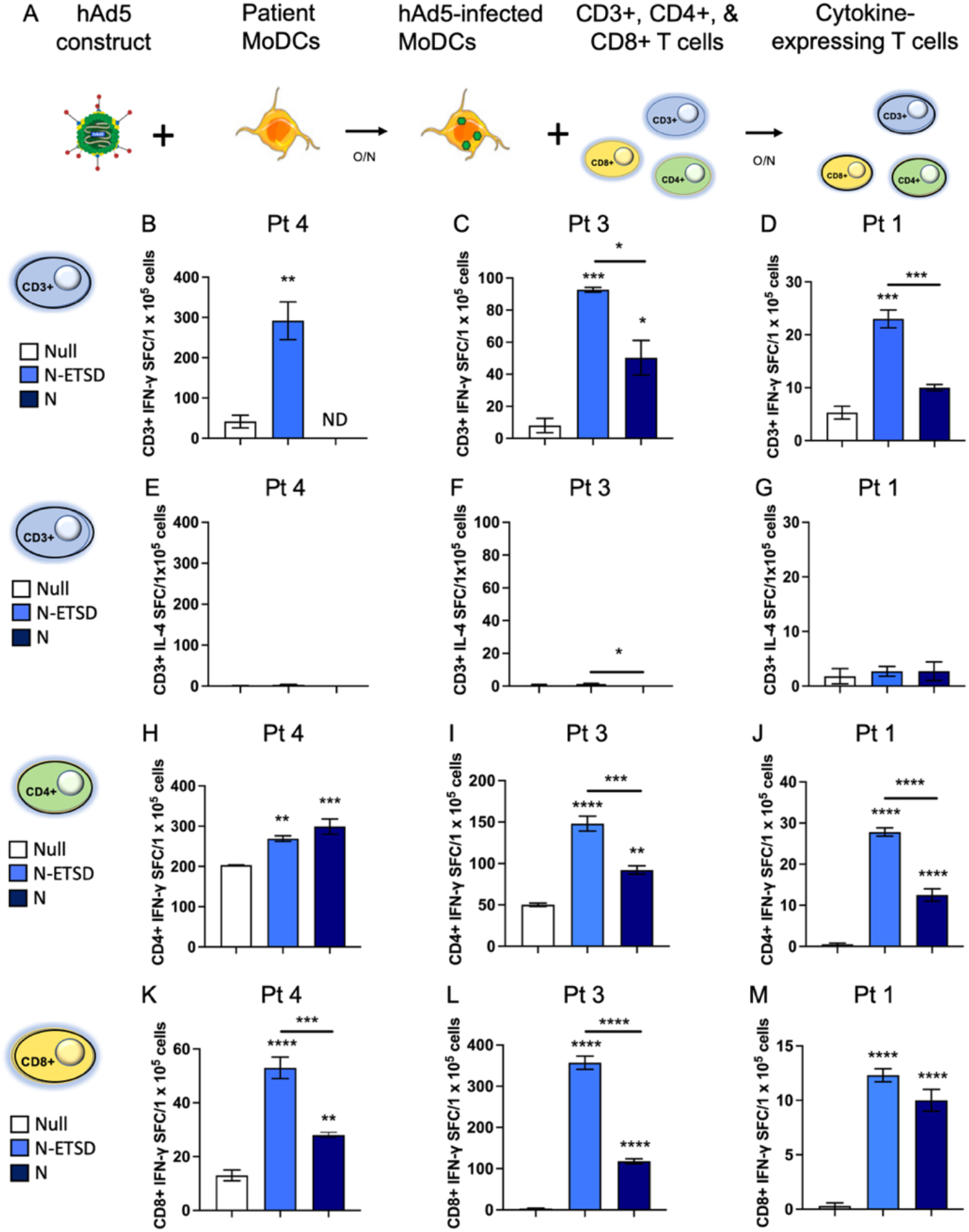
Previously SARS-CoV-2 infected patient T-cell IFN-γ secretion is greater in response to MoDC-expressed N-ETSD than N and is Th1 dominant. (A) Experimental design. (B-C) IFN-γ and (D-F) secretion by autologous CD3+ T cells in response to hAd5 N-ETSD-/N-/Null-expressing MoDCs for 3 previously infected patient. Same scale used for IFN-γ and IL-4 for each patient (B/E; C/F; D/G). IFN-γ secretion by CD4+ (H-J) and CD8+ (K-M) T cells in response to hAd5-N-ETSD/N/Null are shown. Statistical analysis performed using One-way ANOVA and Tukey’s post-hoc multiple comparison analysis, where *p<=0.05; **p<0.01, ***p<0.001 and ****p<0.0001. Comparison to Null shown above bars, comparison between N-ETSD and N indicated with a line. Data graphed as mean and SEM; n = 3-4.

#### CD4+ and CD8+ memory T cells from previously SARS-CoV-2 infected patients recall nucleocapsid and spike antigens expressed by autologous hAd5-infected MoDCs

To compare CD3+, CD4+, and CD8+ memory T-cell responses to the components of the hAd5 S + N vaccine, MoDCs were infected with hAd5 S-Fusion, hAd5 N-ETSD, or the dual antigen vaccine (Fig. 5A).

**Fig. 5.**
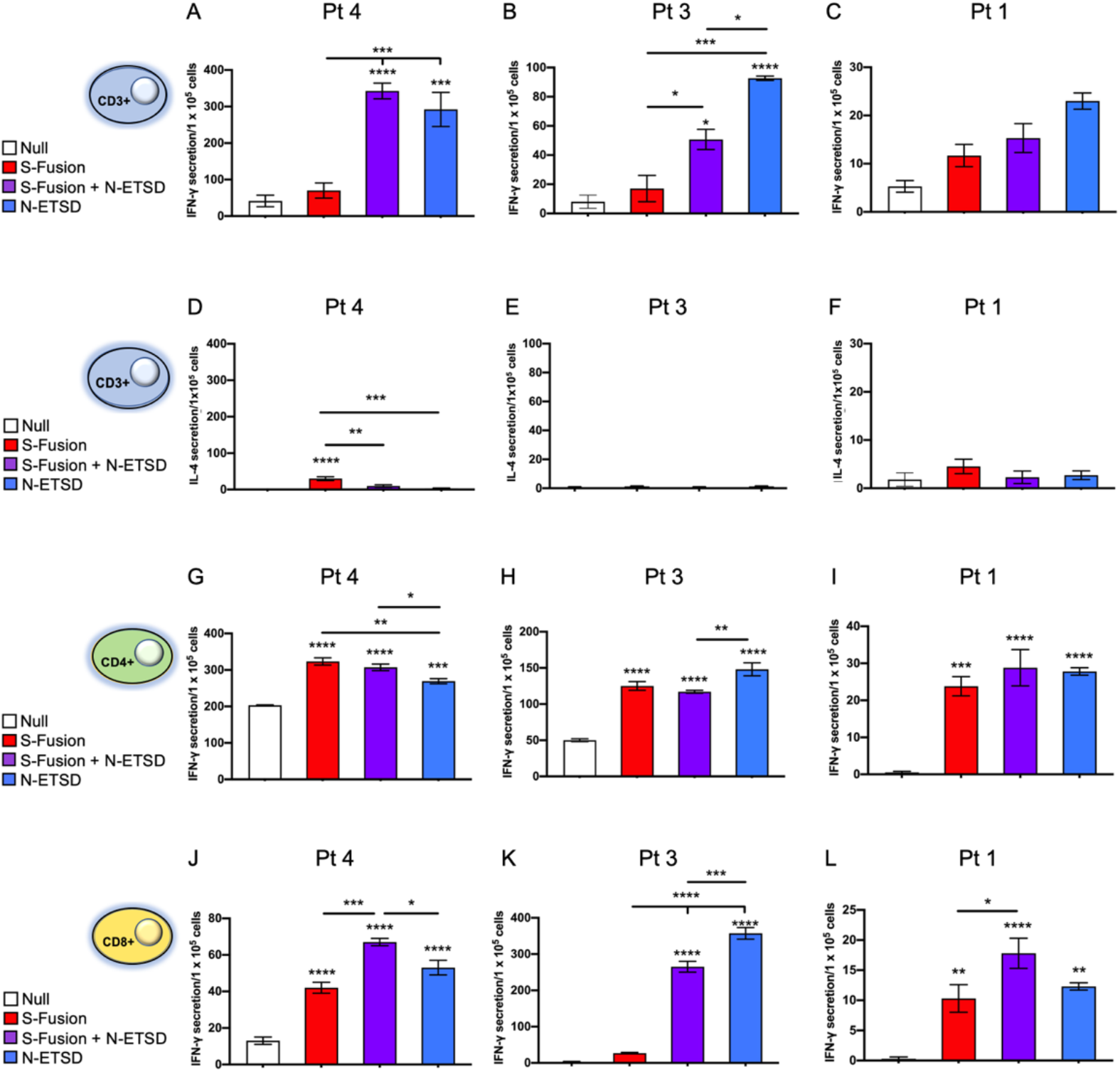
Previously SARS-CoV-2 infected patient T-cell responses to the dual antigen vaccine and its individual components reveal distinct antigen specificity of T-cell populations. (A-C) CD3+ T cell IFN-γ responses for three patients. (D-F) CD3+ T cell IL-4 responses. (G-I) CD4+ IFN-γ responses. (J-L) CD8+ IFN-γ responses. Statistical analysis performed using One-way ANOVA and Tukey’s post-hoc multiple comparison analysis to compare each construct to the Null construct, where *p<0.05; **p ≤ 0.01; ***p ≤ 0.001; and ****p ≤ 0.00001. Comparison to Null only above bars; comparison between antigen-expressing constructs above lines. Data graphed as mean and SEM; n = 3-4.

For unselected CD3+ T cells, IFN-γ responses were similar to S-Fusion + N-ETSD and N-ETSD with responses to S-Fusion being relatively low (Fig. 5A-C). The number of IL-4 secreting T cells in response to any infected MoDC was very low (Fig. 5D-F). Based on the increased expression of S in the dual antigen vaccine compared to monovalent S-Fusion, the increased T-cell responses could be explained by either T cells recognizing increased S or the presence of N. Importantly, these T-cell responses were characterized by a predominance of IFN-γ (Th1) relative to IL-4 (Th2).

CD4+ T cells from all three patients showed significantly greater recognition of all three constructs compared to Null (Fig. 5G-I). While there were greater responses to specific constructs in some individuals, overall the responses to S-Fusion, S-Fusion + N-ETSD and N-ETSD were similar. CD8+ T cells from all three patients recognized the dual antigen and N-ETSD vaccines at a significantly higher level than Null; in only two of three patients did CD8+T cells recognize S-Fusion to a significant degree above Null (Fig. 5J-L). These data indicate that T cells from previously infected SARS-CoV-2 patients have reactivity and immune memory recall to both of the vaccine antigens (S and N) in our vaccine vector.

### T-Cell Responses after a Single Prime Vaccination in a Phase 1 Clinical Trial

For the ELISpot assay results shown here, T cells were isolated from patients in cohort 2 of the Phase 1b study of the ImmunityBio hAd5 COVID-19 ‘T-cell’ vaccine (NCT04591717). These participants were vaccinated subcutaneously with 1 × 10^11^ hAd5 S + N viral particles (VP).

The studies presented here are experimental and all assays and analyses were performed in-house at ImmunityBio, Inc. Formal analyses of the primary and secondary end points are ongoing pending completion of the trial.

#### T cells from vaccinated participants show Th1 dominant responses to S and N by Day 14 after a single prime injection, with a more than 10-fold increase in mean responses to N

T-cell responses to S, N, and SARS-CoV-2 membrane (M) peptides were determined by ELISpot for clinical study participants pre-vaccination (Day 1) and at two time points following the prime injection: Days 14-16 and 21-23.

By Day 14-16, mean T-cell secretion of IFN-γ in response to N peptides increased more than 10-fold compared to pre-vaccination (Day 1); these responses were sustained on Day 21-23 and were significant (Fig. 6A). The changes in T-cell responses to N for individual participants from pre- to post-vaccination are shown in Fig. 6B.

**Fig. 6.**
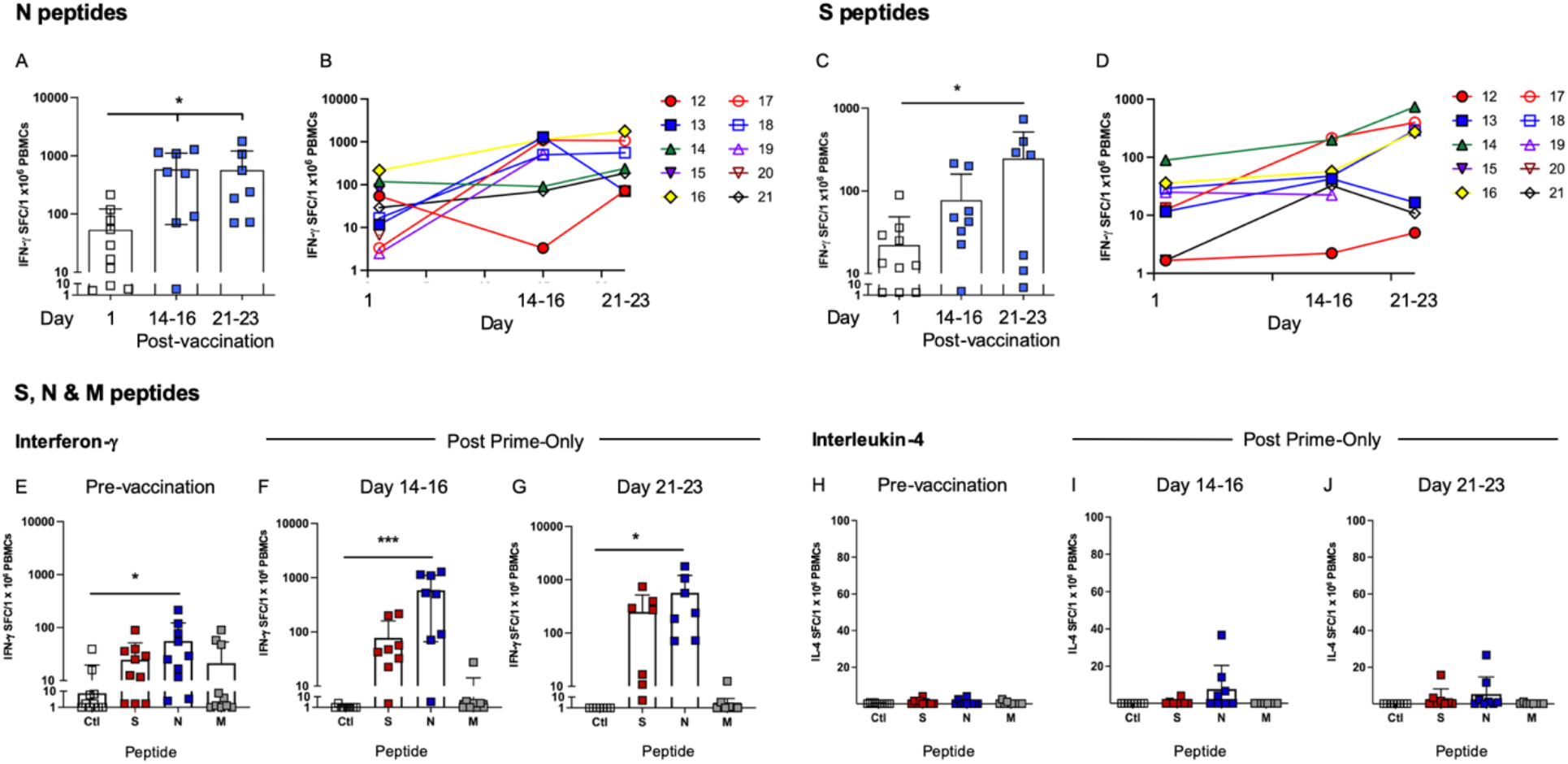
Vaccinated participant T-cell responses to S, N, and M peptides. (A) T-cell interferon-γ (IFN-γ) secretion in response to nucleocapsid (N) peptides is shown for pre-vaccination (Day 1) and post-prime only Day 14-16 and 21-23. (B) Individual participant T-cell responses to N. (C)T-cell IFN-γ secretion in response to spike (S) peptides is shown for pre-vaccination (Day 1) and post-prime only Day 14-16 and 21-23. (D) Individual participant T-cell responses to S. T-cell IFN-γ secretion in response to no stimulation (Ctl), S, N and membrane (M): (E) pre-vaccination and on (F) Day 14-16 and (G) 21-23 post-prime only; (H-J) interleukin-4 (IL-4) secretion. Statistical analysis performed using One-way ANOVA and Sidak’s post-hoc analysis to compare pre-to post-vaccination (A, C) or peptide stimulation to Ctl (E-J), where *p, 0.05, **p ≤ 0.01, and ***p ≤ 0.001. Data graphed as mean and SD; baseline n = 9, Day 14-16 n = 7, Day 21-23 n = 7.

T-cell secretion of IFN-γ in response to S peptides also increased post-prime only vaccination, with the mean increasing more than 3-fold by Day 14-16 and more than 10-fold by Day 21-23, when differences were significant (Fig. 6C). T-cell responses to S for individual participants are shown in Fig. 6D.

When responses to S, N and SARS-CoV-2 membrane (M) peptides are compared, the greatest responses are seen to N peptides (Fig. 6F and G). As expected, there were very low/no responses to M peptide, not delivered in the vaccine. Importantly, IL-4 secretion in response to peptide stimulation was very low, indicating all responses to vaccine antigens were highly Th1 dominant.

#### T-cell responses to S and N antigens generated from hAd5 prime vaccination alone were equivalent to those from previously SARS-CoV-2 infected patients

As shown in Figure 7, T-cell responses to both N and S for previously SARS-CoV-2 infected patients (Pt) and hAd5 S + N vaccinated participants (IB Vac) on Day 21-23 were comparable. For responses to N, both Pt and IB Vac were ∼40-fold greater than Unex; and for responses to S, both Pt and IB Vac were >15-fold higher than Unex. As expected, T cells from previously SARS-CoV-2 infected patients, but not vaccinated participants, reacted to M due to its presence in the live virus. The M antigen is not present in the vaccine.

**Fig. 7.**
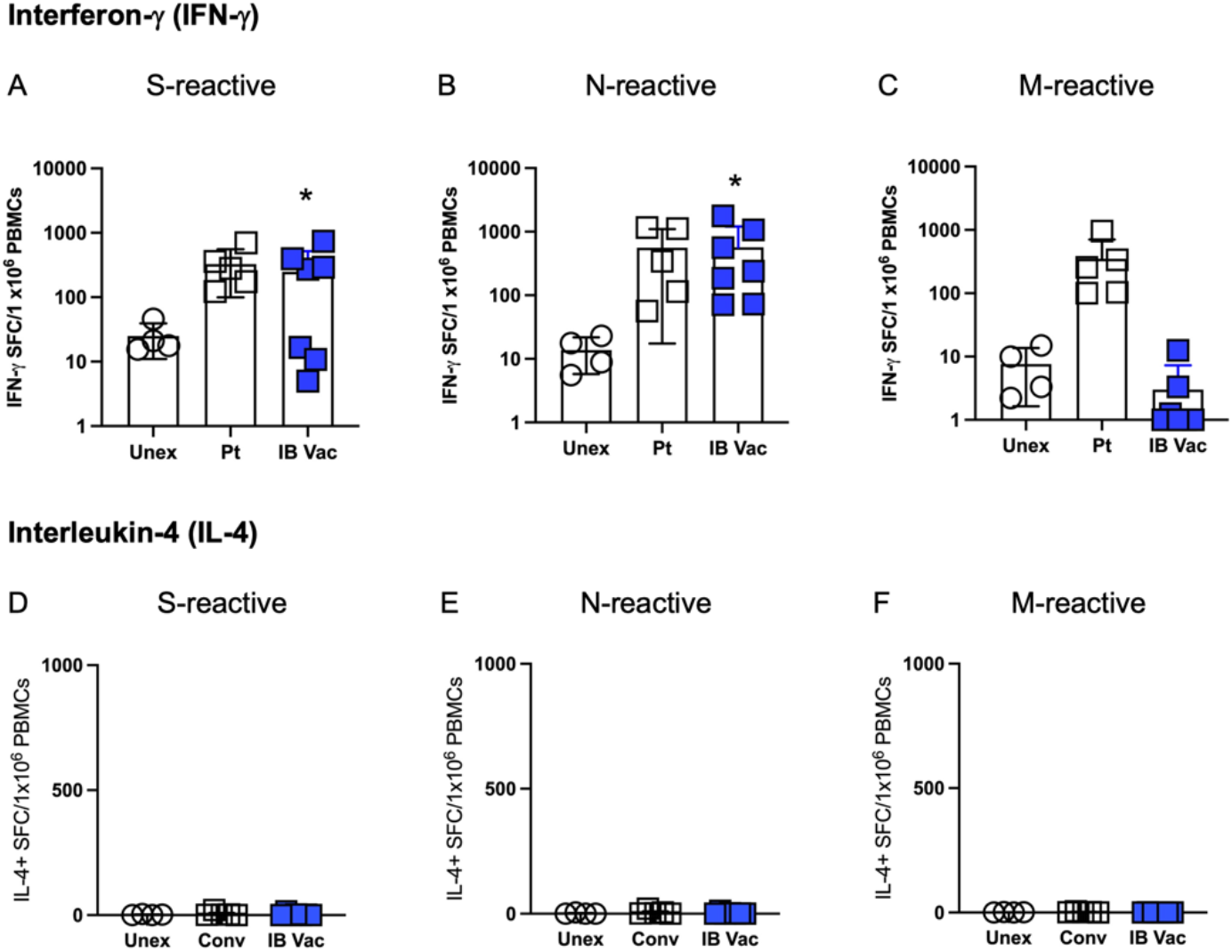
T-cells from vaccinated and previously SARS-CoV-2 infected individuals show similar responses to S and N. T-cell secretion of interferon-γ (IFN-γ) in response to (A) N, (B) S, and (C) M as determined by ELISpot shown for SARS-CoV-2 unexposed (Unex) and previously infected (Pt) individuals as compared to participants receiving a prime vaccination alone (IB Vac) from Day 21-23. Secretion of interleukin-4 (IL-4) in response to (D) N, (E) S, and (F) M. Statistical analysis performed using a Wilcoxon signed-rank test where *p < 0.05. Data graphed as mean and SEM; Unex n = 4, Pt n = 5, IB vac n = 7.

#### T cells from hAd5 S + N vaccinated participants show equivalent reactivity to S wild type and variant peptides

To determine if vaccinated participant T-cell responses seen to S wildtype (WT) peptide pools after only prime vaccination are sustained in response to S variant peptide pools, these responses were assessed by ELISpot to pools comprising the variants B.1.1.7 (H0069δ, V0070δ, Y0144δ, N0501Y, A0570D, D0614G, P0681H, T0716I, S0982A, and D1118H mutations), B.1.429 (S0013I, W0152C, L0452R, and D0614G mutations), P.1/B.1.1.28 clade (L0018F, T0020N, P0026S, D0138Y, R0190S, K0417T, E0484K, N0501Y, D0614G, H0655Y, T1027I, and V1176F mutations), and B.1.351 (D0080A, D0215G, L0242-, A0243-, L0244-, K0417N, E0484K, N0501Y, D614G, and A0701V mutations).

As shown in Figure 8A, for all 3 vaccinated participants from which sufficient PBMCs were available for assessment, IFN-γ secretion in response to the S WT sequence used in the hAd5 S + N vaccine and the mutant S peptide pools was very similar. When graphed together after normalization of T-cell IFN-γ secretion in response to media/S variants to S WT (S WT = 1.0) for each individual, there are no statistical differences between responses to S WT and any variant, while the difference between media and S WT was highly significant.

**Fig. 8.**
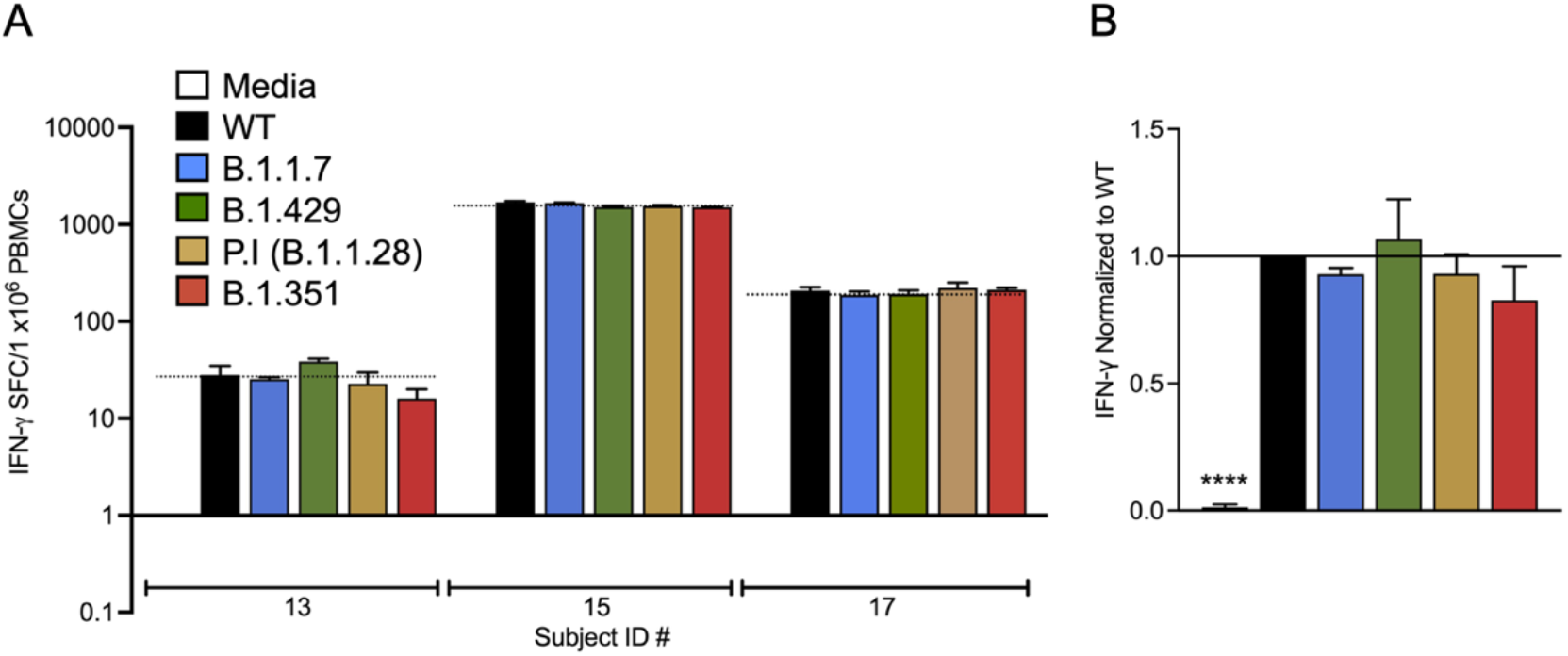
T cells from vaccinated participants show similar responses to S wildtype (WT) and S variants. (A) T-cell secretion of interferon-γ (IFN-γ) in response to medium only (media), S WT peptides or B.1.1.7 S, B.1.429, P.1 (B.1.1.28 clade), and B.1.351 variant peptides as determined by ELISpot is shown for 3 vaccinated participants after a single injection of the vaccine. (B) IFN-γ secretion normalized to S WT for the 3 vaccinated participants combined. Statistical analysis performed using One-Way ANOVA with Dunnett’s post-hoc analysis comparing media and variant peptide pools to the S-WT peptide pool, where *p < 0.05 and ****p ≤ 0.0001; n = 3. Data graphed as the mean plus SEM.

## DISCUSSION

The emergence and rapid spread of SARS-CoV-2 variants is raising concern that first generation monovalent wild-type S-targeting vaccines may eventually show reduced protection against COVID-19. In addition to updating S sequences to include variants in current vaccines, alternate or complimentary vaccine strategies include enhanced focus on eliciting protective T-cell responses and addition of other immunogenic structural proteins, such as M and/or N. Evidence indicates that virtually all COVID-19 convalescents develop IFN-γ-secreting CD4^+^ and/or CD8^+^ T-cell responses not only against S, but also against the M and N proteins of SARS-CoV-2.^31^ Significant T-cell reactivity in COVID-19 convalescents has also been observed against a wide breadth of other SARS-CoV-2 proteins, particularly nsp6 and ORF3a.^28,31,51^ Collectively, the total magnitude of the T-cell response against non-S proteins such as N appears at least equal to or possibly greater than the T-cell response against S in COVID-19 convalescents.^28,31,51^ These data suggest that T-cell responses (when present) elicited by monovalent S-targeting vaccines designed to generate humoral responses likely provide limited cell-mediated protection.

Here we show that our strategy of including N-ETSD, confirmed to be directed to the endo/lysosomal compartment with S optimized for cell-surface display ^1,52^ results in recognition of the N-ETSD and S-Fusion + N-ETSD antigen(s) by previously SARS-CoV-2 patient T cells and in generation of S- and N-reactive T cells in participants receiving the hAd5 S-Fusion + N-ETSD vaccine. The enhancement of T-cell responses by the addition of N in this vaccine confers a greater likelihood of sustained protection against variants as our T-cell epitope HLA binding prediction analysis suggests.

The report of Redd *et al*. ^7^ provides further support for the merits of T-cell protection against variants. In their study, the authors assessed reactivity of CD8+ T cells from thirty COVID-19 convalescent individuals to forty-five different mutations and concluded that virtually all CD8+ T cell responses to SARS-CoV-2 antigens should recognize the variants. The data presented here align with their findings and provide evidence that the hAd5 S-Fusion + N-ETSD vaccine has the potential to provide cellular immunity against SARS-CoV-2 variants.

Our dual-antigen hAd5 S + N vaccine, which is predicted to provide expanded T-cell protection, may have utility as a ‘universal’ booster. Not only would boosting with this vaccine likely increase cell-mediated protection for individuals who have already received a first-generation S-only vaccine, because the N protein is highly conserved among coronaviruses,^53^ the hAd5-S-Fusion + N-ETSD vaccine might provide protection against viruses similar to SARS-CoV-2 that may arise.

The hAd5 S-Fusion + N-ETSD COVID-19 T-cell vaccine is currently undergoing clinical testing with delivery by a variety of routes, including sublingual, SC, intranasal, and a thermally-stable oral tablet as a prime/boost to provide protection against SARS-CoV-2. We anticipate our testing its use as a universal boost for current first generation vaccines in the near future.

## Data Availability

All data are reported in the manuscript

## ACKNOWLEDGEMENTS

We thank the volunteers participating in our Phase I COVID-19 vaccine clinical trial and Deborah Fridman, Director of Clinical Research at Hoag Hospital (Irvine/Newport Beach, CA). We also thank Phil Yang of ImmunityBio for his ongoing coordination of project updates for the studies described herein.

## SUPPLEMENTARY MATERIALS for

**Prime hAd5 Spike + Nucleocapsid Vaccination Induces Ten-Fold Increases in Mean T-Cell Responses in Phase 1 Subjects that are Comparable COVID-19 Convalescents and Sustained Against Spike Variants**

## Supplementary Materials

### Methods

#### Quantification of anti-SARS-CoV-2 spike antibodies in plasma

The presence of anti-SARS-CoV-2 spike antigen antibodies in previously infected patient plasma used in studies here was validated. IgG against SARS-CoV-2 spike was detected in plasma using ELISA. Briefly, EIA/RIA plates were coated with a solution of purified recombinant SARS-CoV-2-derived Spike protein (1μg/ml, S-Fusion. ImmunityBio, Inc.) suspended in coating buffer (0.05 M carbonate-bicarbonate, pH 9.6) and incubated overnight at 4°C. Plates were washed three times with TPBS solution (PBS + 0.05% Tween 20). Blocking solution (2% non-fat milk in TPBS) was added and incubated at room temperature (RT, 1h). Serial dilutions of plasma were prepared in 1% non-fat milk in TPBS. Plates were washed as described above and serial dilutions of plasma were added to the plate and incubated at RT (1h). Plates were washed three times. Goat anti-Human IgG (H+L) HRP-conjugated secondary antibody (1:6000 dilution) prepared in 1% non-fat milk/TPBS was added and incubated at RT (1h). Plates were washed three times. Substrate (3,3’,5,5’-tetramethylbenzidine (TMB)) was added to each well and incubated at RT (10min). The reaction was stopped by addition of sulfuric acid (1N H2SO4). The optical density (450 nm) was measured by a Synergy 2 (BioTek Instruments, Inc.) plate reader. Data were analyzed using Prism 8 (GraphPad Software, LLC).

#### cPass™ neutralizing antibody detection

The GenScript cPass™ (https://www.genscript.com/cpass-sars-cov-2-neutralization-antibody-detection-Kit.html) kit for detection of neutralizing antibodies was used according to the manufacturer’s instructions.^41^ The kit detects circulating neutralizing antibodies against SARS-CoV-2 that block the interaction between the S RBD with the ACE2 cell surface receptor. It is suitable for all antibody isotypes.

To evaluate the levels of anti-SARS-CoV-2 neutralizing antibodies in plasma, dilutions of plasma were incubated with horseradish peroxidase-conjugated spike RBD (37°C, 30min). The RBD-plasma mixture was added to a microtiter plate coated with ACE2 and incubated at 37°C (15 min). Plates were washed and substrate (TMB) added at room temperature (15 min). The reaction was stopped and plates read on a plate reader at 450 nm.

#### Live virus assay of plasma neutralization of infection

The ability of previously SARS-CoV-2 infected patient plasma used in studies here to neutralize SARS-CoV-2 infection *in vitro* was also validated. All aspects of the assay utilizing virus were performed in a BSL3 containment facility according to the ISMMS Conventional Biocontainment Facility SOPs for SARS-CoV-2 cell culture studies. Vero E6 kidney epithelial cells from *Cercopithecus aethiops* (ATCC CRL-1586) were plated (20,000 cells/well) in a 96-well format and 24 hours later, cells were incubated with antibodies or heat inactivated plasma previously serially diluted in 3-fold steps in DMEM containing 2% FBS, 1% NEAAs, and 1% Pen-Strep; the diluted samples were mixed 1:1 with SARS-CoV-2 in DMEM containing 2% FBS, 1% NEAAs, and 1% Pen-Strep at 10,000 TCID 50/mL for 1 hr at 37°C, 5% CO2. This incubation did not include cells to allow for neutralizing activity to occur prior to infection. For detection of neutralization, the virus/sample mixture (120 μL) was transferred to the Vero E6 cells and incubated (48 hours) before fixation with 4% PFA. Each well received virus (60 μl) or an infectious dose of 600 TCID50. Control wells, including six wells on each plate for no virus and virus-only controls, were used. The percent neutralization was calculated as 100-((sample of interest-[average of “no virus”])/[average of “virus only”])*100) with a stain for CoV-2 Np imaged on a Celigo Imaging Cytometer (Nexcelom Bioscience).

#### Transfection of HEK 293T cells and detection of plasma antibody binding

To assess binding of plasma from a previously infected patient to antigens expressed by hAd5 S-Fusion and hAd5 S-Fusion + N-ETSD vaccines, plasma was incubated with construct-transfected HEK 293T cells and binding determined by flow cytometry. HEK 293T cells (2.5 × 10^5^ cells/well in 24 well plates) were grown in DMEM (Gibco) with 10% FBS and 1X PSA (100 units/mL penicillin, 100 μg/mL streptomycin, 0.25 μg/mL Amphotericin B) at 37°C. Cells were either left untransfected or were transfected with 0.5 µg of S-Fusion or S-Fusion + N-ETSD hAd5 plasmid DNA using a JetPrime transfection reagent (Polyplus) according to the manufacturer’s instructions. Twenty-four hours later, cells were incubated for 30 min. with previously infected patient or healthy (unexposed, UnEx) plasma that had been serially diluted 10-fold for 30 min. Plasma IgG was labeled using a goat anti-human IgG-phycoerythrin conjugated and labeled cells were acquired using the Thermo-Fisher Attune NxT flow cytometer and analyzed using FlowJo Software to determine Mean Fluorescence Intensity (MFI) values of both the untransfected and transfected cells. Results were graphed as the difference in MFI between untransfected and transfected cells for the S-Fusion and S-Fusion + N-ETSD constructs over the plasma dilutions.

### Results

#### Previously SARS-CoV-2-infected patient plasma confirmed to have neutralizing anti-S antibodies

For the studies described below, plasma samples were collected from four individuals convalescing from SARS-CoV-2 infection as confirmed by antibody assays and patient history as described Methods. The presence of anti-Spike IgG, and of neutralizing antibodies by both a surrogate SARS-CoV-1 neutralization assay ^41^ and live virus assays, were confirmed in all patient samples (Suppl. Fig. S1). Samples were also collected from four virus-naïve individuals and were used as controls.

**Fig. S1.**
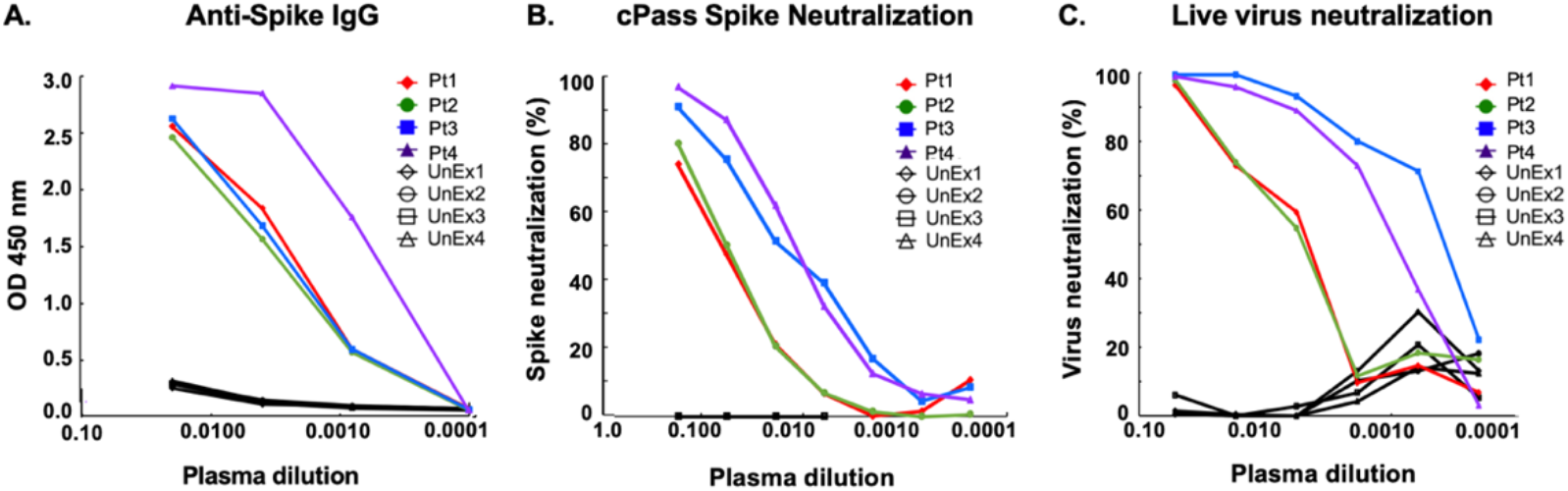
Plasma samples from patients (Pt) previously infected with SARS-CoV-2 contain neutralizing antibodies. (A) The levels of anti-spike IgG as determined by OD at 450 nm are shown. (B) All four Pt plasma samples contained antibodies that blocked RBD-ACE2 binding and (C) contained SARS-CoV-2 neutralizing antibodies.

#### Previously SARS-CoV-2-infected patient plasma binds to vaccine-transfected HEK-293T cells

In additional studies to validate immune responses to SARS-CoV-2 antigens, the binding of previously SARS-CoV-2 infected patient and virus-naïve control individual plasma to human embryonic kidney (HEK) 293T cells transfected with either hAd5 S-Fusion alone or hAd5 S-Fusion + N-ETSD was assessed (Suppl. Fig. S2). This binding reflects the presence of antibodies in plasma that recognize antigens expressed by the hAd5 vectored vaccines. Quantification of histograms showed little or no binding of virus-naïve plasma antibodies to cells expressing either construct, and the highest binding of plasma antibodies from a previously SARS-CoV-2 infected patient to cells expressing the dual antigen S-Fusion + N-ETSD construct (Suppl. Fig. S2R).

**Fig. S2.**
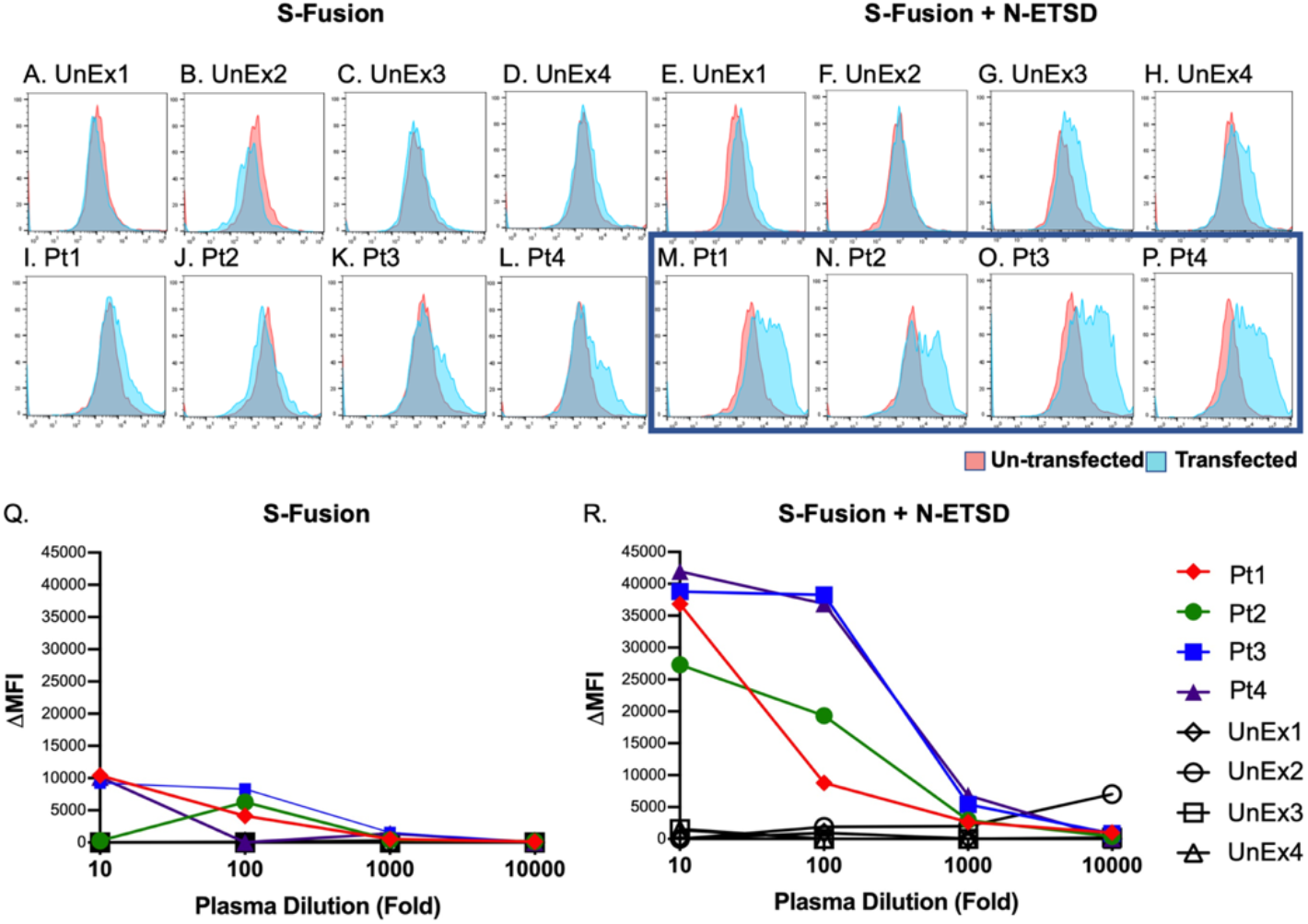
Plasma from previously SARS-CoV-2 infected patients shows greater binding to S-Fusion + N-ETSD surface antigens compared to S-Fusion alone after transfection of HEK 293T cells. HEK 293T cells that were either uninfected (pink) or infected (blue) with S-Fusion alone or bivalent S-Fusion + N-ETSD were exposed to plasma from either unexposed (UnEx) controls or previously infected SARS-CoV-2 patients (Pt). Flow histograms for (A-D) UnEx with S-Fusion; (E-H) UnEx with S-Fusion + N-ETSD; (I-L) Pt with S-Fusion; and (M-P) Pt with S-Fusion + N-ETSD, all n = 4, are shown. When the ΔMFI (difference in binding to transfected versus untransfected cells) is graphed, it is apparent that compared to (Q) S-Fusion, (R) binding of Pt plasma to S-Fusion + N-ETSD transfected cells is much higher and that plasma from UnEx individuals shows little binding.

#### Previously SARS-CoV-2-infected patient T cells are activated by S and N peptide pools

The reactivities of T cells from the previously SARS-CoV-2-infected patients was confirmed by incubation with S1 (containing the spike receptor binding domain), S2 and N peptides pools (Fig. S3A). The specific reactivities of CD4+ (Fig. S3B and C) and CD8+ T cells was then determined, showing CD8+ T cells were more selectively activated by the N peptide pool.

**Fig. S3.**
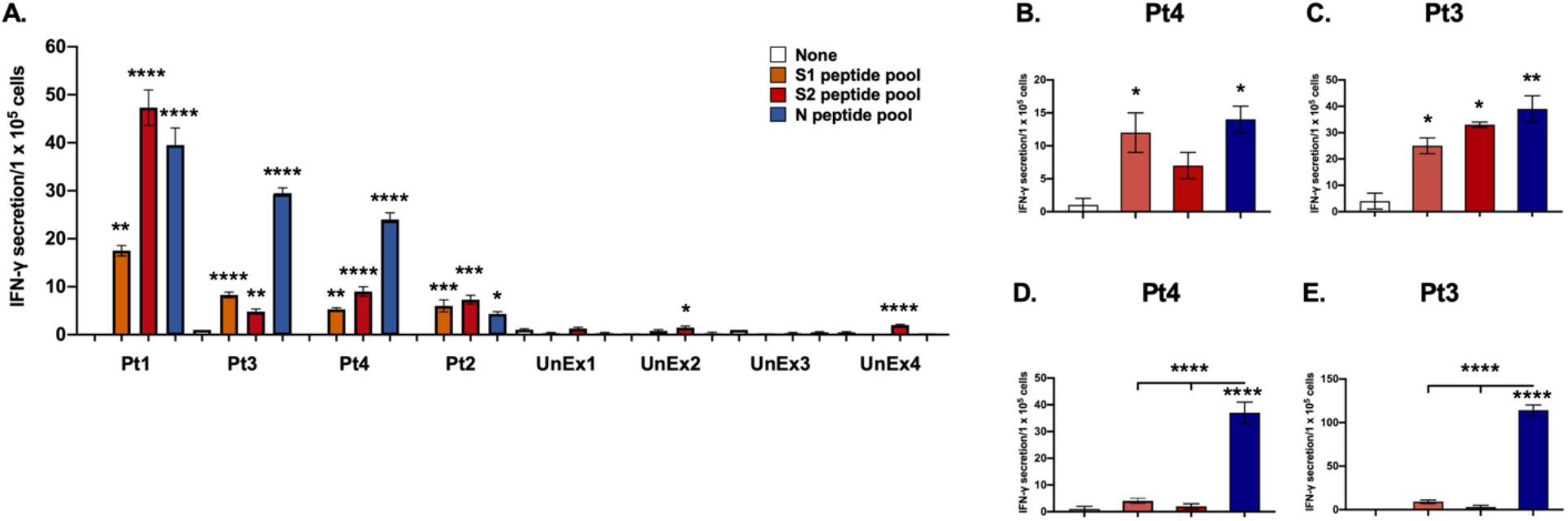
T-cell responses to MoDCs pulsed with SARS-CoV-2 peptides. Unselected T cells from all four previously SARS-CoV-2 infected patients (Pt) show significant IFN-γ responses to S1, S2, and N peptide pool-pulsed MoDCs as compared to ‘none’. T cells from virus-naïve (unexposed, UnEx) control individuals showed far lower responses. Statistical analysis performed using One-way ANOVA and Tukey’s post-hoc analysis for samples from each patient compared only to ‘none’ where * p<= 0.05, **p<0.01, ***p<0.001, and ****p<0.0001. Data graphed as the mean and SEM; n = 3-4. Patient MoDCs pulsed with SARS-CoV-2 peptide mixes overnight were incubated with autologous CD4+ (B, Pt 4; C, Pt 3) or CD8+ (D, Pt 4; E, Pt 3) T cells. IFN-γ levels were determined by ELISpot. Statistical analysis performed using One-way ANOVA and Dunnett’s post-hoc multiple comparison analysis to compare each peptide pool to Veh (shown above the bar) or between peptide pools (above line) where *p<0.05, **p<0.01 and ****p<0.00001. Data graphed as mean and SEM; n = 3.

